# Gaze fixation stability is a transdiagnostic marker of major psychiatric disorders: A high-density family-based study

**DOI:** 10.1101/2025.06.23.25330103

**Authors:** Swarna Buddha Nayok, Vanteemar S Sreeraj, Sonika Nichenmetla, Harleen Chhabra, Pavithra Jayasankar, Srinivas Balachander, Bharath Holla, Biju Viswanath, Vivek Benegal, YC Janardhan Reddy, Mathew Varghese, Sanjeev Jain, ADBS-CBM Consortium, John P John, Ganesan Venkatasubramanian

## Abstract

**Background:** Eye movement tracking non-invasively captures subtle cognitive and neural differences. Fixation stability, the ability to maintain steady visual fixation, may serve as a transdiagnostic marker in major psychiatric disorders. This study evaluates fixation stability as a potential transdiagnostic endophenotype in families with multiple affected individuals.

**Methods:** Monocular eye tracking data was recorded using infrared cameras while participants fixed their gaze on a stimulus in trials with and without distractors. The fixation stability (FS) performance was compared across 449 individuals affected with major psychiatric disorders (26 Alzheimer’s Dementia (AD), 89 schizophrenia (SCZ), 116 bipolar disorder (BD), 98 obsessive-compulsive disorder (OCD), and 120 substance-use disorder (SUD)), 442 unaffected first degree relatives (FDRs) along with 145 healthy controls (HC). FS performance was compared across groups using a linear mixed effects model controlling for familiality, age and sex.

**Result:** Affected individuals performed significantly poorly (fixation frequency(F=6.37, p_cor_=0.003), median fixation duration(F=4.79, p_cor_=0.009), saccade frequency(F=7.74, p_cor_<0.001), mean saccade amplitude(F=4.92, p_cor_=0.009), mean scanpath length(F=6.83, p_cor_=0.003)) in the trials with distractors when compared to FDRs and HC. The performance of FDRs and HC did not differ significantly from that of the other. Furthermore, in a cross-diagnostic comparison, impaired performance was observed only in SCZ and BD, with both performing significantly worse than SUD, OCD, and HC.

**Conclusions:** FS performance was impaired in major psychiatric disorders compared to FDRs and HC. Instead of an endophenotype, FS measures serve as illness markers. SCZ and BD showed the greatest deficits, highlighting the strong impact of psychotic conditions on visuoperceptual processing.

## 1. Introduction

Eye movements influence various human behaviours, from fundamental visual perception for survival and everyday decision-making to complex cognitive and emotional processes underlying human social structure (1). Even simple eye movements associated with gazing at a target represent basic but complex visuoperceptual processing (2,3). Apart from involving the retina, the optic tract and the occipital lobe, visuoperceptual processing requires optimal functioning of several brain regions like the superior colliculus (SC), geniculate bodies, medio-posterior cerebellum, midbrain and frontotemporal lobes (3).

Abnormal or inadequate visuoperceptual processes are found in several psychiatric disorders. Deficits in smooth pursuit and saccadic movements are seen in schizophrenia (4). Similar deficits are found even in simple free-gaze viewing tasks in schizophrenia. Theoretically, symptoms of schizophrenia have been linked to impairments in adaptation and optimisation of context-dependent responses to visual stimuli (gain control) and facilitation of interacting neural responses to achieve co-activation of various related processes (integration) (5,6). Further, poor visual motion perception and deficits in smooth pursuit and vergence have been found in both bipolar disorder and schizophrenia, suggestive of shared neurophysiological insults (7,8). Early visual perception, assessed by visual evoked potentials in the occipital fields, is found to be defective in obsessive-compulsive disorder (OCD) as well (9). Eye movements are linked to reward processing and become important in addiction (10). Indeed, perceptuomotor and visuospatial abnormalities related to visual attention load and contextual processing are dysregulated in addiction (11,12). Visual perceptual organisation, such as contour detection and integration, is deficient in Alzheimer’s dementia (13,14). Several aspects of visual processing and integration are abnormal in other psychiatric disorders like depression (15) and anxiety (16,17) and in children with autism (18).

As eye movement and visuoperceptual processing abnormalities are found in several psychiatric disorders, the etiopathological, clinical and therapeutic implications of such abnormalities are being studied. For example, eye movement abnormalities have been proposed to be diagnostic and therapeutic biomarkers in schizophrenia (4,19,20), OCD (21) and neurodegenerative disorders (22). Machine learning algorithms, which typically include gaze fixation and saccades (4,22,23), can be employed to advance the knowledge of the potential biomarkers further.

The fixation stability task (FS) is a simple eye movement task that measures the ability to gaze at a fixed target and informs us regarding basic visuoperceptual processes (2,3). Visual stability depends on optimal integration of several neuronal processes, like attention, reafference in visual field motion, proprioception, and corollary discharge (24,25). Such synchronous processes involve several brain regions like the fovea, SC, and medio-posterior cerebellum. It may specifically involve omnipause neurons that are responsible for controlling saccadic movements (3). FS is proposed to serve as a visual function biomarker (26). Evaluating such basic processes may inform us regarding the common underlying abnormalities in psychiatric disorders and serve as a potential etiological, diagnostic, clinical, and prognostic marker. Recently, performance in FS tasks integrated with other eye movement measures has been shown to distinguish schizophrenia from healthy controls (23). However, research also indicates that such deficits may not be exclusive to schizophrenia and may be found in bipolar disorder (27), OCD (28,29), and dementia (30). Therefore, FS deficits in several psychiatric disorders may serve as a transdiagnostic marker that perhaps implies shared pathophysiology. As underlying early visual processing deficits span across psychiatric disorders, such transdiagnostic measures may help identify individuals vulnerable to developing major psychiatric disorders.

Further, FS measures have been proposed to serve as an endophenotype marker of schizophrenia (31,32), but the findings are inconsistent (27,32). There is also a suggestion that FS deficits could be transdiagnostic (23). Thus, the study evaluates fixation stability in a large group of patients with major psychiatric disorders (schizophrenia (SCZ), bipolar disorder (BD), OCD, alcohol dependence/substance use disorder (SUD) and Alzheimer’s dementia (AD)), their FDRs and healthy controls (HC). We hypothesised FS deficits to be a transdiagnostic endophenotype, with individuals affected with major psychiatric disorders and their unaffected FDRs to perform poorer than HC. Further, we hypothesized that the FS deficits may present differentially across the major psychiatric disorders, with deficits particularly prominent in those with psychotic disorders.

## 2. Methods and Materials

### 2.1 Participants

This study included participants from the Accelerator Program for Discovery in Brain Disorders using Stem Cells - Centre for Brain and Mind (ADBS-CBM) (33,34). The ADBS-CBM is an ongoing family-based study where patients with any of the five major psychiatric disorders (SCZ, BP, OCD, SUD, AD), their unaffected FDRs, and population HC are evaluated longitudinally. Each family in this study has a minimum of two FDRs with one of the five major psychiatric disorders, along with their unaffected FDRs. Individuals who do not have any lifetime psychiatric disorder and without any of the above psychiatric disorders in their FDRs are included as HC. Further, the clinical diagnosis of the affected individual was also evaluated independently by two psychiatrists. The study was approved by the Institutional Ethics Committee. All participants provided written informed consent.

### 2.2 Eye tracking methodology and FS task

Detailed clinical evaluation, including transdiagnostic Clinical Global Impressions (CGI) severity scores and physical examination, preceded eye movement tracking. Any individuals with major ophthalmological/ neurological/ medical conditions or having uncorrected refractory errors were excluded from this analysis.

Eye tracking task, data acquisition and analysis were done as per Sreeraj et al. (35). Ocular dominance was evaluated using the Dolman method, and the dominant eye was tracked using infra-red cameras (EyeLink® 1000 eye tracker, SR Research, Canada) in an illuminance-controlled room at a 1000 Hz sample rate. The participants performed the FS task while sitting comfortably with their chin and headrest to reduce head movement. In this task, a 0.5^0^ central circular yellow target would appear for 5 seconds on the black background on a 22-inch flat-screen monitor (FuzHion, Viewsonic, 120 Hz refresh rate, 1680×1050 pixels resolution) (36). The participants were asked to gaze at this target, not look elsewhere. In some trials, within those 5 seconds, another identical target (called a distractor) would appear at either 1.43^0^ or 2.86^0^ distance on either of the sides in the horizontal plane. The distractor would appear in the presence of the target, and only one distractor would appear in the trial. Participants were instructed at the start of the task to ignore any such distractors. There were two blocks with five trials in each. The first trial of each block was without distractors; therefore, there were two trials without and eight trials with distractors. The trials within each block were presented pseudorandomly. Trials without distractors are considered easier to perform than trials with distractors (23,24,37).

### 2.3 Eye movement data processing

After converting the EyeLink eye-tracking data to the generic ASCII format, a custom-made script based on the PyGaze toolbox was used for further processing (38). Fixation events less than 40 ms, nearing the boundary of the screen, outside the screen, saccades with amplitude <0.1° or >100°, saccades with duration <10 ms or >300 ms, saccades starting or ending outside the screen’s edge, blink period with adjacent ±50ms periods data were excluded (23,39).

Fixation frequency, average fixation duration (ms), median fixation duration (ms), saccade frequency, average saccadic amplitude (°), and scan path length (°) from each trial were measured and averaged over the trials with and without a distractor. Higher fixation and saccade frequencies, average saccadic amplitude, scan path length, and lower median fixation duration indicate poor FS performance.

### 2.4 Statistical analysis

Statistical analysis was done using Statistical Package for the Social Sciences (IBM Corp., Armonk, N.Y., USA). Socio-demographical details were compared using Analysis of Variance (ANOVA) for continuous variables and Chi-square for categorical variables. FS measures between the affected individuals, FDRs and HC were compared using serial linear mixed-effect models. Further, these measures were compared between the five psychiatric disorders and HC. Age and sex were used as fixed-effect variables for all the models. As ADBS-CBM is a family study, several individuals belonging to either of the groups may belong to the same family. To control for familial effects, individuals within a family were used as random effect variables. Benjamini-Hochberg method of family-wise error (FWE) correction was used to avoid type-1 error at a p-value of 0.05 (p_cor_) (40). Post-hoc Bonferroni tests were used during pairwise comparisons between the groups. Pearson’s correlation was used to assess the relationship between the affected group’s CGI scores and FS measures, controlling for age.

## 3 Results

### 3.1 Group description

FS performance was analysed for 449 affected, 451 FDRs (from about 379 multiplex families) and 145 HC (N=1036). Within the affected patients, 26 had AD, 89 had SCZ, 116 had BD, 98 had OCD, and 120 had SUD. The sociodemographic details are given in **Table 1** and **Table 2**. Age and gender differed significantly across the groups. Within the affected group, age positively correlated with total illness duration and negatively with years of education. Total illness duration negatively correlated with years of education. CGI severity did not correlate with age, age at onset, illness duration, or years of education.

**Table 1:** Sociodemographic details of affected, FDR and HC (N=1036)

| Table 1: Sociodemographic details of affected, FDR and HC (N=1036) |  |  |  |  |  |  |  |  |
| --- | --- | --- | --- | --- | --- | --- | --- | --- |
| Sociodemographic variables |  | Affected (n=449) | FDR (n=442) | HC (n=145) | Comparison between groups |  |  |  |
|  |  |  |  |  | F/X<sup>2</sup> | df | p | η<sup>2</sup><sub>p</sub>/Cramer V |
| Age (Mean years±SD) (n=1036) |  | 39.91±13.83 | 41.37(15.2) | 32.41(14.56) | 21.08* | 2 | <0.001 | 0.04 |
| % of males |  | 61.92 | 45.02 | 51.72 | 25.70** | 2 | <0.001 | 0.16 |
| Years of education (Mean years±SD) (n=1031) |  | 8.81±5.43 | 10.01±5.73 | 13.56±4.3 | 40.46* | 2 | <0.001 | 0.07 |
| Occupation, % (n=978) | Employed | 50.59 | 55.53 | 48.23 | 129.91** | 10 | <0.001 | 0.26 |
|  | Homemaker | 20.43 | 23.56 | 6.38 |  |  |  |  |
|  | Retired | 4.28 | 4.33 | 6.38 |  |  |  |  |
|  | Student | 8.08 | 12.5 | 36.17 |  |  |  |  |
|  | Unemployed | 16.63 | 4.09 | 2.13 |  |  |  |  |
| df: degrees of freedom, η<sup>2</sup><sub>p</sub>: partial eta squared, FDR: first-degree relatives, HC: healthy control, SD: Standard deviation, *ANOVA, **Chi-square |  |  |  |  |  |  |  |  |

**Table 2:** Sociodemographic details of five psychiatric disorders (N=449)

| **Table 2: Sociodemographic details of five psychiatric disorders (N=449)** |  |  |  |  |  |  |  |  |  |  |
| --- | --- | --- | --- | --- | --- | --- | --- | --- | --- | --- |
| Sociodemographic variables |  | AD (n=26) | SCZ (n=89) | BD (n=116) | OCD (n=98) | SUD (n=120) | Comparison between groups |  |  |  |
| | | | | | | | F/X<sup>2</sup> | df | p | $\eta^2_p$/<br>Cramer V |
| Age (Mean years±SD)<br>(n=449) |  | 64.69±7.57 | 39.2±11.78 | 38.41±14.04 | 35.80±13.7 | 39.86±10.52 | 29.4* | 4 | <0.001 | 0.2 |
| % of males |  | 45.83 | 45.45 | 49.56 | 57.29 | 92.44 | 67.88** | 4 | <0.001 | 0.39 |
| Years of education<br>(Mean years±SD)<br>(n=445) |  | 11.89±5.85 | 8.55±5.56 | 8.71±5.03 | 9.56±6.23 | 8.09±4.68 | 3.17* | 4 | 0.01 | 0.03 |
| Occupation, % (n=417) | Employed | 20.83 | 31.7 | 50.9 | 41.76 | 78.18 | 152.07** | 16 | <0.001 | 0.3 |
|  | Homemaker | 25 | 28.05 | 23.63 | 25.27 | 6.36 |  |  |  |  |
|  | Retired | 37.51 | 3.7 | 1.81 | 2.20 | 2.72 |  |  |  |  |
|  | Student | - | 6.09 | 13 | 17.58 | - |  |  |  |  |
|  | Unemployed | 16.67 | 32.93q | 13.0 | 13.19 | 12.73 |  |  |  |  |
| CGI severity scores<br>(Mean±SD)<sup>#</sup> (n=383) |  | 3.52±0.77 | 2.83±1.35 | 2.08±1.26 | 3.82±1.12 | 4.26±1.24 | 49.53*** | 4 | <0.001 | 0.35 |
| CGI severity % (CGI as a<br>categorical variable)<br>(n=383) | Normal | - | 23.61 | 48.42 | 3.49 | 3.92 | 179.29** | 20 | <0.001 | 0.34 |
|  | Borderline ill | 4.17 | 16.67 | 16.84 | 6.98 | 8.82 |  |  |  |  |
|  | Mildly ill | 50 | 22.22 | 18.95 | 27.91 | 9.8 |  |  |  |  |
|  | Moderately ill | 33.33 | 30.56 | 11.58 | 29.07 | 20.59 |  |  |  |  |
|  | Markedly ill | 12.5 | 4.17 | 3.16 | 30.23 | 49.02 |  |  |  |  |
|  | Severely ill | - | 2.78 | 1.05 | 2.33 | 7.84 |  |  |  |  |
| Total duration of illness<br>(Mean years±SD)<br>(n=352) |  | 2.12±2.54 | 11.74±10.14 | 14.95±11.26 | 13.44±11.18 | 15.72±10.11 | 8.64* | 4 | <0.001 | 0.09 |
| Age at onset (Mean<br>years±SD) (n=351) |  | 61.55±7.03 | 27.19±8.32 | 23.36±9.26 | 20.79±7.71 | 24.18±8.55 | 108.32* | 4 | <0.001 | 0.56 |

df: degrees of freedom, $\eta^2_p$ : partial eta squared, AD: Alzheimer's Disease, BD: Bipolar Disorder, OCD: Obsessive-Compulsive Disorder, SCZ: Schizophrenia, SD: Standard deviation, SE: Standard error, SUD: Substance Use Disorder, \*ANOVA, \*\*Chi-square, \*\*\*Analysis of covariance, # significant effect of age ( $p<0.001$ )

### 3.2 Group differences: Affected vs FDRs vs HC

The FS measures in the trials without distractors were comparable across the three groups (**Supplementary Table S1, Figure 1**). However, with the introduction of distractors, affected individuals performed significantly poorer in FS compared to their FDRs and HCs (**Table 3**, **Figure 1**). The unaffected FDRs performed similarly to HCs. There was a significant effect of age and gender, with younger individuals and males having better FS (p<0.05) (**Supplementary Table S2**). Estimates of family as the random effect parameter are given in **Supplementary Table S3.**

**Figure 1:**
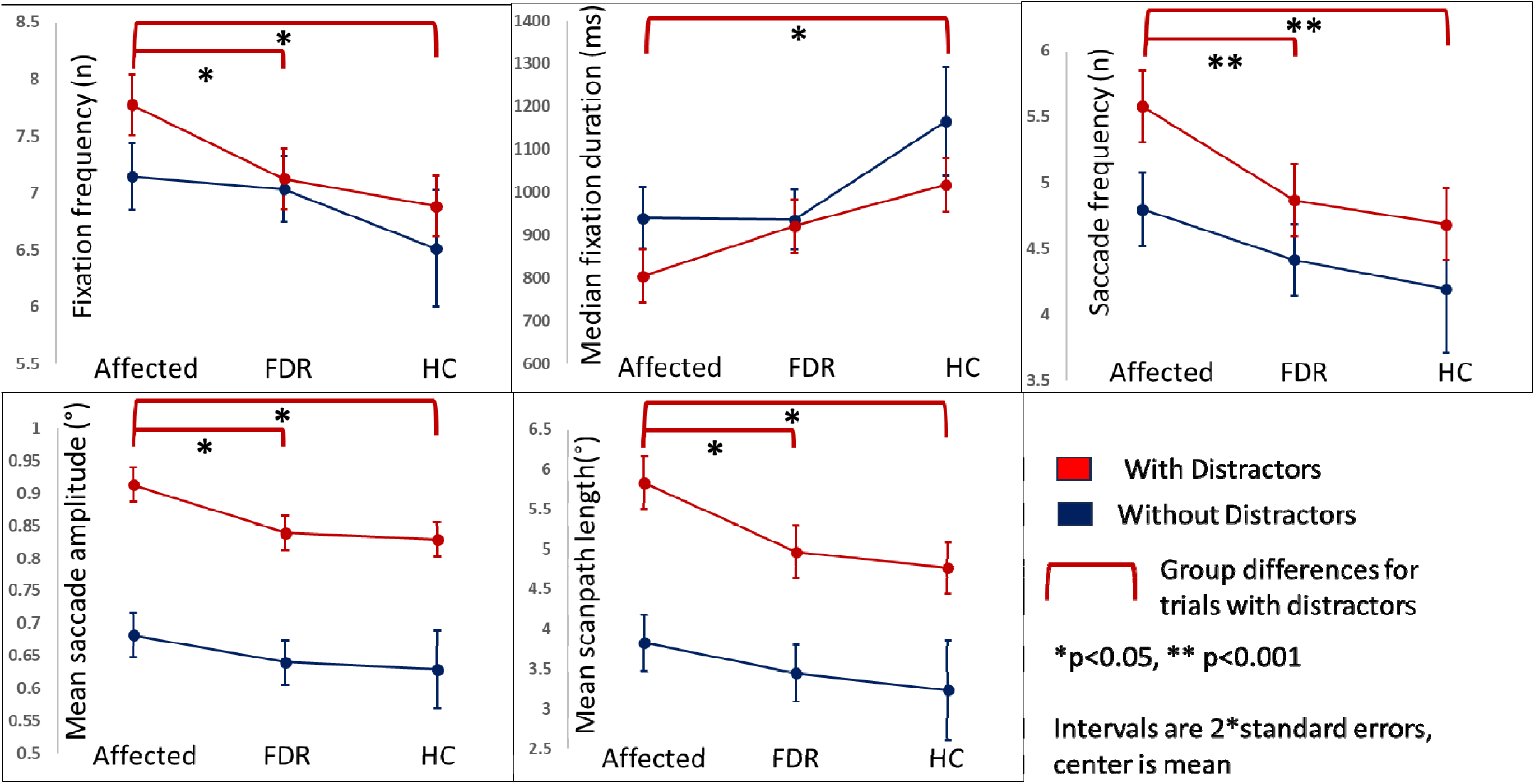
Fixation stability measures across the groups (Affected, FDR, HC) for trials with (blue) and without (red) distractors. FDR: first-degree relatives, HC: healthy control

**Table 3:** Group comparisons between Affected, FDR and HC for trials with distractors.

|  |  |  |  |  |  |  |
| --- | --- | --- | --- | --- | --- | --- |
| **FS measures** | **Estimated marginal means** |  |  | **Type III Tests of Fixed Effects** |  |  |
|  | **Affected (n=449)** | **FDR (n=442)** | **HC (n=145)** | **F** | **$p_{cor}$** | **Posthoc** |
|  | **Mean (SE)** | **Mean (SE)** | **Mean (SE)** |  |  |  |
| Fixation frequency | 7.73 (0.15) | 7.14 (0.15) | 6.88 (0.26) | 6.37 | 0.003 | Affected>HC, FDR |
| Median fixation duration | 812.18 (37.61) | 914.30 (37.14) | 1018.59 (63.67) | 4.79 | 0.009 | Affected<HC |
| Saccade frequency | 5.53 (0.15) | 4.89 (0.14) | 4.67 (0.24) | 7.74 | <0.001 | Affected>HC, FDR |
| Mean saccade amplitude | 0.91 (0.02) | 0.84 (0.02) | 0.83 (0.03) | 4.92 | 0.009 | Affected>FDR |
| Mean scanpath length | 5.77 (0.19) | 5.0 (0.18) | 4.74 (0.32) | 6.83 | 0.003 | Affected>HC, FDR |
| FS: fixation stability, FDR: first-degree relatives, HC: healthy control, SCZ: schizophrenia, SE: standard error, $p_{cor}$: Adjusted p-value | | | | | | |

As cognitive deficits are a defining feature of AD, a sensitivity analysis excluding AD was performed to note similar results showing significant poor performance in the affected group (**Supplementary Table S4 & S5**). The effect of age persisted to be significant despite removing patients with AD (**Supplementary Table S4**). To understand whether the effect of diagnosis is a product of differences in age across the groups, we compared the current model with two other models – one without age and another without diagnosis. We found that for most of the FS measures, the current model (fixed effects: age, sex, diagnosis, random effect: family) had the best model fitting suggesting the significant contributions of diagnosis and age independent of each other (**Supplementary Table S6**).

### 3.3 Disorder specific effects

There were no significant group differences in the trials without distractors after controlling for age and gender (**Supplementary Table S7, Figure 2**). However, only SCZ and BD had impaired FS compared to HC in trials with distractors. SUD, OCD and AD had no significant difference in FS compared to HC. Indeed, the performance of SUD and OCD was significantly better than SZ and BD (**Table 4**, **Figure 2**). Effects of age and sex are given in **Supplementary Table S8**. Sensitivity analysis after removing AD yielded similar results (**Supplementary Table S9 and S10**). Again, as done in section 3.2, we compared the current model with those without age and diagnosis separately, and found that for most FS measures, the current model showed the best fit, suggesting the significant contributions of diagnosis and age independent of each other (**Supplementary Table S11**).

**Figure 2:**
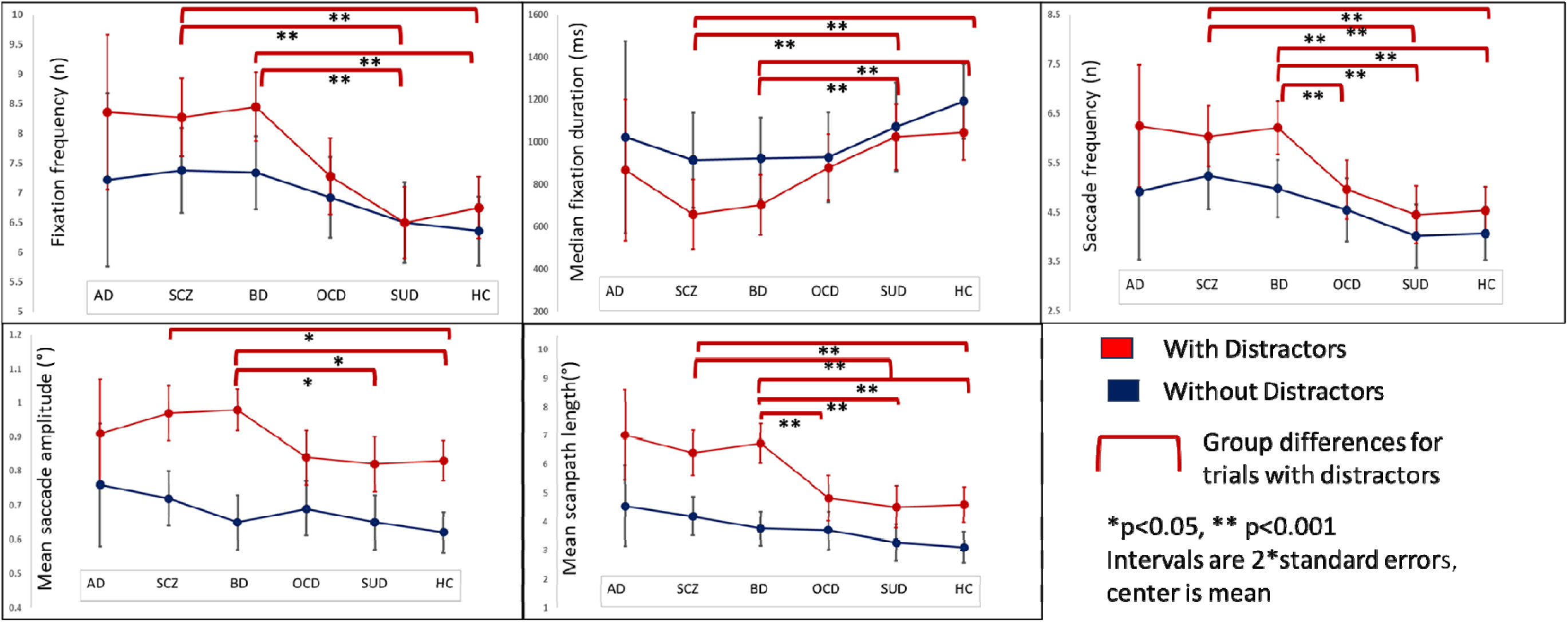
Fixation stability measures across the groups (AD, SCZ, BD, OCD, SUD, HC) for trials with (blue) and without (red) distractors. AD: Alzheimer’s Disease, BD: Bipolar Disorder, FDR: first-degree relatives, HC: healthy control, OCD: Obsessive-Compulsive Disorder, SCZ: Schizophrenia, SUD: Substance Use Disorder

**Table 4:** Group comparisons between AD, SCZ, BD, OCD, SUD and HC for trials with distractors.

|  |  |  |  |  |  |  |  |  |  |
| --- | --- | --- | --- | --- | --- | --- | --- | --- | --- |
| **FS measures** | **Estimated marginal means** |  |  |  |  |  | **Type III Tests of Fixed Effects** |  |  |
|  | **AD (n=26)**<br>**Mean (SE)** | **SCZ (n=89)**<br>**Mean (SE)** | **BD (n=116)**<br>**Mean (SE)** | **OCD (n=98)**<br>**Mean (SE)** | **SUD (n=120)**<br>**Mean (SE)** | **HC (n=145)**<br>**Mean (SE)** | **F** | **p<sub>cor</sub>** | **Posthoc** |
| Fixation frequency | 8.34(0.67) | 8.20(0.33) | 8.43(0.30) | 7.31(0.32) | 6.45(0.30) | 6.74(0.26) | 7.33 | <0.001 | SCZ, BD>HC, SUD |
| Median fixation duration | 865.04 (163.3) | 667.84 (82.12) | 704.67 (72.84) | 875.30 (79.80) | 1035.32 (76.23) | 1043.09 (65.13) | 4.53 | <0.001 | SCZ, BD<HC, SUD |
| Saccade frequency | 6.20(0.61) | 5.97(0.31) | 6.2(0.27) | 4.98(0.3) | 4.4(0.29) | 4.54(0.25) | 7.34 | <0.001 | SCZ>HC,SUD;<br>BD>HC,SUD,OCD |
| Mean saccade amplitude | 0.92(0.08) | 0.97(0.04) | 0.98(0.03) | 0.84(0.04) | 0.82(0.04) | 0.82(0.03) | 4.1 | 0.001 | SCZ>HC;<br>BD>HC,SUD |
| Mean scanpath length | 7.01(0.8) | 6.31(0.39) | 6.71(0.35) | 4.76(0.38) | 4.46(0.36) | 4.59(0.31) | 7.8 | <0.001 | SCZ,BD>HC,SUD,<br>OCD |
| AD: Alzheimer's dementia, BD: bipolar disorder, FS: fixation stability, HC: healthy control, p<sub>cor</sub> : Adjusted p-value, OCD: obsessive compulsive disorder, SCZ: schizophrenia, SE: standard error, SUD: substance (alcohol) use disorder |  |  |  |  |  |  |  |  |  |

### 3.4 Clinical Severity and FS

Although most patients were clinically stable, the CGI score was significantly different across the diagnoses (**Table 2**), with SUD having the highest mean CGI severity score, followed by AD, OCD, SCZ and BD. There were no significant correlations between CGI severity scores and any FS performance metric of trials with distractors. Significant negative correlations with CGI severity scores were found in fixation and saccade frequencies and mean scanpath length within the affected group (**Supplementary Table S12**). However, no significant correlations were found when individual disorder groups were compared. Similarly, age of onset did not significantly correlate with FS measures when comparing individual disorder groups.

### 3.5 Assessing endophenotypic trend within SCZ and BD

As described in section 3.2, FDR did not seem to have significantly lower FS performance scores than HC, suggesting that perhaps these FS measures are not endophenotypes.

However, these measures were significantly poorer in those affected with SCZ and BD. To determine whether these measures presented an endophenotypic pattern, affected with only SCZ and BD, FDRs of those with SCZ and/or BD and HC were compared as three groups. The results showed similar patterns; those with SCZ and BD performed significantly poorly compared to FDRs and HC, with no significant differences between FDRs and HC (**Supplementary Table S13**). FS measures seem to have no endophenotypic trend even when FDRs of SCZ and BD are compared with HC and SCZ/BD.

## Discussion

The current study compared the FS across the three groups (affected, unaffected FDRs, and HC) to evaluate its value as an endophenotype in this sample of high-density families. The study was conducted in families with a high density of psychiatric illness so that it would be more sensitive in identifying the biological signals. However, FDRs performed as well as HC despite poor performance by their affected relatives, as detected in most of the FS measures, suggesting that FS measures are probably illness-related markers and not endophenotypes. The analysis further indicated segregation of the FS abnormalities in patients with SZ and BD but not others, suggesting that these measures are perhaps not transdiagnostic (for SCZ/BD, OCD, SUD and AD) in nature.

### 4.1 Distractors bring out poor performance in major psychiatric disorders

Poorer fixation stability was apparent only when distractors were presented along with the target stimuli. Affected individuals were unable to control their gaze away from the distractors. This resulted in the induction of more saccades, less time staying with the target stimuli and, hence, an increase in the frequency of fixations (23,24,37). On getting distracted, non-affected individuals (both familial and population controls) could rectify soon to stay longer with the target stimuli. However, affected individuals had to look at the distractors, thereby increasing the length of the saccade amplitude and the length of the path used for visual scanning.

It is not clear whether attention is a prerequisite for all the eye movements and whether attention and awareness of the eye movements are dissociable from each other (41,42). Previous studies propose that the subject may be unaware of the loss of attention while performing eye movement tasks. A “certain amount” of attentional change may be required for awareness of attentional change. This change in attention, but perhaps not awareness, may be brought by the presence of distractors. Subsequently, the FS tasks may show poor performance, pointing towards poorer attention, while the subject may still perceive to be focussed on the central stimuli (43,44). The effects of distractors may vary based on their position and relevance and may differ in different eye-tracking tasks. In the current task, the distractors seem to induce poor FS performance even in HC, especially in saccade frequency, amplitudes, and scanpath length. The poor performance was further prominent in those affected with psychiatric disorders.

Further, previous studies suggest that shared spatial representations may connect fixation and attention (24). At a “rudimentary” level, FS functions adequately with either enough attention or due to the presence of a visual target for fixation. However, spatial representation may be disturbed when the visual target is present, especially when visual distractors also appear beside the initial target. When visual distractors are present, more attention may be required for the eyes to remain fixated on the target. The HC seems to have overcome these hurdles and adequately fixed their eyes on the target. The deficits in spatial allocation, target fixation, and poor attention may all contribute to poor performance in those with major psychiatric disorders, especially in those with SCZ and BD (4).

Eye movement measures such as fixation and saccades are strongly influenced by age in the general population (45,46). With increasing age, the movements are slower and poorer, becoming evident by the third decade of life. Previous research indicates that performance in fixation tasks often correlates with age, as in the case of the current study (47). Such age-related effects are more significant when tasks are often repeated, perhaps due to a cumulative effect (48). Age-related changes, task difficulty level, and the disease process may play a complex role in eye movements, requiring further evaluation. The findings of distractor’s influence were robust for the five measures of the FS even after controlling for age and sex and correction for multiple comparisons.

As AD consists of individuals of a higher age, a sensitivity analysis was performed to remove AD from the group of affected individuals. Due to neuro-ocular degenerative changes, AD may result in poor performance in FS tasks (49). However, the results remained similar, with the affected group showing poor FS performance in several measures. It seems that age may have an independent effect on eye fixation and is not influenced by the disorder. This further increases the complexities of interpreting FS measures, as age may continue to have differential effects on other neurophysiological measures in major psychiatric disorders (50). On the contrary, it may be easier to compare FS measures across psychiatric disorders presenting at various age groups (51). Indeed, this may help overcome the previous hurdles of comparing neurophysiological measures due to age differences in several psychiatric disorders, making eye-tracking tasks better suitable for comparative studies.

### 4.2 FS as an endophenotype marker of major psychiatric disorders

To be considered an endophenotypic marker, the measure should be present differentially in the affected, their FDRs and HC (52,53). The current study shows that FS measures are predominantly poor only in the affected group. The absence of a significant difference in these measures between FDR and HC goes against considering these as endophenotype markers. These measures may show basic psychiatric illness processes, such as visuoperceptual processing, which are markers of the illness process. Perhaps the ability of the distractors to compromise the fixation of the eye may serve as a potential illness marker of major psychiatric disorders. This may help to differentiate psychiatric illness transdiagnostically, as the FS measures differed significantly between the affected and the HC/FDRs (52). Moreover, FS measures may change with the severity of the psychiatric illness and act as an illness severity marker. Most patients in this current study were in relatively stable conditions, and therefore, relationship between the FS performance and illness severity could not be evaluated. Also, as most of the affected group were on psychiatric medications, FS measures may also be evaluated in the light of the treatment process. If treatments do affect FS measures, these can be further tested as prognostic markers (54).

### 4.3 FS measures in specific disorders

While FS performance was significantly poorer in the affected group when compared to HC and FDR, pointing towards its transdiagnostic utility, further analyses showed that the poor FS performance was significant in those with SCZ and BD. When each major psychiatric disorder is compared to HC, SCZ and BD perform poorly not only from HC but also from SUD and OCD in some measures. Such differences may be traced to intrinsic eye movement deficits in psychotic disorders, although studies have not consistently found differences between SCZ and BD with HC (4,27,55,56). While such deficits are also argued to be specific endophenotypes, their association with clinical states has also been noted (56). FS performance may get impaired even when acute symptoms are minimal (57). However, inconsistent findings are also present, perhaps owing to the medication’s effects on performance (27,58). Medication status remains a confounding factor in the current study. FS abnormalities may point towards subtle but important visual processing deficits, which negatively affect bottom-up processing and may become more prominent during florid psychotic states (59,60). In SCZ and BD, these deficits are accentuated by related pathophysiology, as eye movement disorders are common in psychotic spectrum disorders (31,61).

In the current study, OCD and SUD seemed to perform the best in FS measures among the other major psychiatric disorders. Eye-movement-related abnormalities in OCD and SUD have been attributed to cognitive control, which is perhaps measured better by anti-saccade tasks rather than FS (29,55,62). The current study evaluated whether even basic visual processing is a deficit in OCD and SUD. Although the FS performances are lower than HC, there is no significant difference between OCD, SUD and HC when evaluated as individual disorders. Moreover, differential performance in these tasks seen in SCZ/BD and OCD/SUD may point towards the requirement of different cognitive faculties in the disease process. Comparative studies are required to understand such findings.

This study combines a trans-diagnostic approach with basic physiological measures such as eye movements in a large sample of patients with a major psychiatric illness and HCs. We controlled for familiality, age, and sex using a mixed model approach to robustly show that those with psychiatric disorders find it difficult to fixate on a point, even for 5 seconds, when there are distractors around it. However, we had a relatively smaller sample in AD for comparative analysis across different psychiatric disorders. While the affected group was in stable clinical condition, their CGI scores differed significantly within the groups, which can affect the FS measures. Lifetime severity scores were not present.

## 5. Conclusion

This study demonstrates the utility of FS as a transdiagnostic state marker of visual processing deficits underlying major psychiatric disorders. Additionally, differences in fixation stability during eye movement tasks had the highest deficits in SCZ and BD,indicating a substantial association of psychotic conditions on fundamental visuoperceptual processing. The current study emphasizes that FS performance may be an illness marker of SCZ and BD, and is perhaps not endophenotypic. Future studies should elaborate on the usefulness of the various FS measures and whether these have differential effects on the disorder type and clinical stages.

## Declaration of generative AI and AI-assisted technologies in the writing process

During the preparation of this work, the author(s) used ChatGPT and Grammarly to improve the language and grammar. After using this tool/service, the author(s) reviewed and edited the content as needed and take(s) full responsibility for the content of the publication.

## Author statement

The current study was conceptualized by VSS, HC, SB, BH, BV, VB, YCJR, MV, SJ, JPJ & GV. Methodology was done by VSS, SN, HC. Data acquisition was done by SN, HC, VSS under the supervision of all stated authors. Data curation and formal analysis was done by VSS, SBN, SN, HC. Original draft was written by VSS & SBN with inputs from SB, PJ & BH under the supervision of GV, JPJ, BV, YCJR & SJ. The draft was reviewed by all the authors. The project administration and funding acquisition was by BV, GV, JPJ, PM, YCJR, SJ along with other investigators of the ADBS-CBM consortium.

All the authors have contributed to this study and have reviewed and approved the final version of the manuscript.

## Data Availability

All data produced in the present work are contained in the manuscript

