## Supplementary Materials for "Gaze fixation stability is a transdiagnostic marker of major psychiatric disorders: A high-density family-based study"

| **Supplementary Table S1: Group comparisons between affected, FDR and HC for trials without distractors** | | | | | |
| --- | --- | --- | --- | --- | --- |
|  | **Estimated marginal means** | | | **Type III Tests of Fixed Effects** | |
| **FS measures** | **Affected (n=449)** | **FDR (n=442)** | **HC (n=145)** |  |  |
|  | **Mean (SE)** | **Mean (SE)** | **Mean (SE)** | **F** | **p_cor_** |
| Fixation frequency | 7.14(0.16) | 7.03(0.16) | 6.51(0.28) | 1.94 | 0.19 |
| Median fixation duration | 934.82(47.09) | 939.47(47.25) | 1166.23(81.74) | 3.33 | 0.18 |
| Saccade frequency | 4.78(0.15) | 4.43(0.15) | 4.20(0.26) | 2.43 | 0.19 |
| Mean saccade amplitude | 0.68(0.02) | 0.65(0.02) | 0.63(0.03) | 1.11 | 0.33 |
| Mean scanpath length | 3.78(0.16) | 3.48(0.16) | 3.23(0.27) | 1.92 | 0.19 |
| FS: fixation stability, FDR: first-degree relatives, HC: healthy control, SCZ: schizophrenia, SE: standard error, p_cor_ : Adjusted p-value | | | | | |

| **Supplementary Table S2: Effects of age and sex on FS measures for trials without and with distractors when compared between three groups** **(Affected, FDR, HC)** | | | | | | | | | | |
| --- | --- | --- | --- | --- | --- | --- | --- | --- | --- | --- |
|  | **Fixation frequency** | | **Median duration of fixation** | | **Saccade frequency** | | **Mean saccade amplitude** | | **Mean scanpath length** | |
|  | **F** | **p_cor_** | **F** | **p_cor_** | **F** | **p_cor_** | **F** | **p_cor_** | **F** | **p_cor_** |
| **For trials without distractors** | | | | | | | | | | |
| **Intercept** | 237.208 | <0.001 | 367.294 | <0.001 | 60.264 | <0.001 | 224.851 | <0.001 | 27.236 | <0.001 |
| **Diagnosis** | 1.936 | .19 | 3.323 | .18 | 2.431 | .19 | 1.107 | .33 | 1.918 | .19 |
| **Sex** | 18.046 | <0.001 | 3.529 | .061 | 12.120 | .001 | 6.033 | .014 | 13.020 | <0.001 |
| **Age** | 68.187 | <0.001 | 61.628 | <0.001 | 76.865 | <0.001 | 15.177 | <0.001 | 53.183 | <0.001 |
| **For trials with distractors** | | | | | | | | | | |
| **Intercept** | 286.719 | <0.001 | 469.169 | <0.001 | 78.671 | <0.001 | 468.346 | <0.001 | 45.072 | <0.001 |
| **Diagnosis** | 6.372 | .003 | 4.787 | .009 | 7.737 | <0.001 | 4.921 | .009 | 6.832 | .003 |
| **Sex** | 16.025 | <0.001 | 11.075 | .001 | 11.107 | .001 | .937 | .333 | 7.741 | .006 |
| **Age** | 105.333 | <0.001 | 73.314 | <0.001 | 125.900 | <0.001 | 31.103 | <0.001 | 88.118 | <0.001 |
| FS: fixation stability, FDR: first-degree relatives, HC: healthy control, p_cor_ : Adjusted p-value | | | | | | | | | | |

| **Supplementary Table S3: Estimates of Covariance of family number as random effect for trials without and with distractors when compared between three groups** **(Affected, FDR, HC)** | | | | | | | |
| --- | --- | --- | --- | --- | --- | --- | --- |
| **FS measures** | **Parameter** | **Estimate** | **Std. Error** | **Wald Z** | **p value (uncorrected)** | **95% Confidence Interval** | |
|  |  |  |  |  |  | **Lower Bound** | **Upper Bound** |
| **For trials without distractors** | | | | | | | |
| **Fixation frequency** | Residual | 10.57 | .61 | 17.39 | <0.001 | 9.45 | 11.83 |
|  | Family number (Variance) | .45 | .42 | 1.07 | .284 | .07 | 2.82 |
| **Median duration of fixation** | Residual | 895531.78 | 52106.03 | 17.19 | <0.001 | 799013.86 | 1003708.72 |
|  | Family number (Variance) | 41856.94 | 37020.88 | 1.13 | .258 | 7394.55 | 236931.62 |
| **Mean scanpath length** | Residual | 10.23 | .56 | 18.28 | <0.001 | 9.19 | 11.38 |
|  | Family number (Variance) | .18 | .34 | .52 | .603 | .004 | 7.74 |
| **For trials with distractors** | | | | | | | |
| **Fixation frequency** | Residual | 8.23 | .46 | 17.76 | <0.001 | 7.37 | 9.19 |
|  | Family number (Variance) | 1.05 | .35 | 2.97 | .003 | .54 | 2.03 |
| **Median duration of fixation** | Residual | 513885.28 | 29750.83 | 17.27 | <0.001 | 458761.31 | 575632.84 |
|  | Family number (Variance) | 55002.90 | 22946.11 | 2.4 | .017 | 24281.82 | 124591.92 |
| **Saccade frequency** | Residual | 7.43 | .42 | 17.74 | <0.001 | 6.65 | 8.3 |
|  | Family number (Variance) | .93 | .32 | 2.93 | .003 | .48 | 1.82 |
| **Mean saccade amplitude** | Residual | .12 | .01 | 17.97 | <0.001 | .1 | .13 |
|  | Family number (Variance) | .01 | .01 | 2.51 | .012 | .01 | .03 |
| **Mean scanpath length** | Residual | 12.23 | .69 | 17.84 | <0.001 | 10.96 | 13.65 |
|  | Family number (Variance) | 1.57 | .52 | 3.03 | .002 | .82 | 3.00 |
| FS: fixation stability, FDR: first-degree relatives, HC: healthy control. The final Hessian matrix was not positive definite although all convergence criteria are satisfied for Saccade frequency and Mean saccade amplitude for trials without distractor. | | | | | | | |

| **Supplementary Table S4: Effects of age and sex on FS measures for trials with distractors when compared between three groups without AD** **(Affected without AD, FDR, HC)** | | | | | | | | | | |
| --- | --- | --- | --- | --- | --- | --- | --- | --- | --- | --- |
|  | **Fixation frequency** | | **Median duration of fixation** | | **Saccade frequency** | | **Mean saccade amplitude** | | **Mean scanpath length** | |
|  | **F** | **p_cor_** | **F** | **p_cor_** | **F** | **p_cor_** | **F** | **p_cor_** | **F** | **p_cor_** |
| **Intercept** | 278.241 | <0.001 | 447.194 | <0.001 | 78.380 | <0.001 | 443.019 | <0.001 | 46.687 | <0.001 |
| **Diagnosis** | 5.858 | .006 | 4.893 | .01 | 6.960 | .005 | 4.407 | .01 | 5.666 | .006 |
| **Sex** | 17.352 | <0.001 | 11.413 | .001 | 11.935 | .001 | .892 | .345 | 7.478 | .007 |
| **Age** | 96.417 | <0.001 | 70.047 | <0.001 | 113.972 | <0.001 | 27.610 | <0.001 | 76.932 | <0.001 |
| AD: Alzheimer’s dementia, FS: fixation stability, FDR: first-degree relatives, HC: healthy control, p_cor_ : Adjusted p-value | | | | | | | | | | |

| **Supplementary Table S5: Group comparisons between affected without AD, FDR and HC for trials with distractors** | | | | | | |
| --- | --- | --- | --- | --- | --- | --- |
|  | **Estimated marginal means** | | | **Type III Tests of Fixed Effects** | | |
| **FS measures** | **Affected (n=423)** | **FDR (n=442)** | **HC (n=145)** |  |  |  |
|  | **Mean (SE)** | **Mean (SE)** | **Mean (SE)** | **F** | **p_cor_** | **Posthoc** |
| Fixation frequency | 7.66(0.16) | 7.09(0.15) | 6.83(0.26) | 5.86 | 0.004 | Affected>HC, FDR |
| Median fixation duration | 814.05(38.84) | 928.67(37.96) | 1025.63  (64.11) | 4.89 | 0.009 | Affected<HC |
| Saccade frequency | 5.45(0.15) | 4.84(0.14) | 4.62(0.24) | 6.96 | 0.007 | Affected>HC, FDR |
| Mean saccade amplitude | 0.90(0.02) | 0.83(0.02) | 0.83(0.03) | 4.41 | 0.012 | Affected>FDR |
| Mean scanpath length | 5.65(0.19) | 4.96(0.18) | 4.67(0.31) | 5.67 | 0.005 | Affected>HC, FDR |
| FS: fixation stability, FDR: first-degree relatives, HC: healthy control, SCZ: schizophrenia, SE: standard error, p_cor_ : Adjusted p-value | | | | | | |

| **Supplementary Table S6: Model fit as per Bayesian information criteria scores for trials with and without distractors when compared between three groups** **(Affected, FDR, HC) and between three groups without AD** | | | | | | | | | |
| --- | --- | --- | --- | --- | --- | --- | --- | --- | --- |
| **Affected vs FDR vs HC** | | | | | | | **Affected without AD vs FDR vs HC** | | |
|  | **Trials without distractors** | | | **Trials with distractors** | | | **Trials with distractors** | | |
|  | **Age, sex & diagnosis** | **Age & sex** | **Diagnosis & sex** | **Age, sex & diagnosis** | **Age & sex** | **Diagnosis & sex** | **Age, sex & diagnosis** | **Age & sex** | **Diagnosis & sex** |
| **Fixation frequency** | 5447.56 | 5449.74 | 5505.59 | 5260.70 | 5271.33 | 5352.75 | 5117.52 | 5127.15 | 5201.27 |
| **Median fixation duration** | 17149.92 | 17177.56 | 17213.10 | 16628.66 | 16658.20 | 16702.26 | 16227.61 | 16257.43 | 16298.11 |
| **Saccade frequency** | 5318.84 | 5321.76 | 5384.87 | 5153.28 | 5166.40 | 5263.77 | 5008.32 | 5019.91 | 5108.03 |
| **Mean saccade amplitude** | 1035.67 | 1027.64 | 1038.42 | 841.57 | 840.73 | 859.71 | 832.74 | 830.96 | 847.58 |
| **Mean scanpath length** | 5389.39 | 5391.42 | 5433.14 | 5669.85 | 5682.18 | 5746.64 | 5510.97 | 5521.01 | 5577.39 |
| AD: Alzheimer’s dementia, FS: fixation stability, FDR: first-degree relatives, HC: healthy control | | | | | | |  |  |  |

| **Supplementary Table S7: Group comparisons between AD, SCZ, BD, OCD, SUD and HC for trials without distractors** | | | | | | | | |
| --- | --- | --- | --- | --- | --- | --- | --- | --- |
| **Estimated marginal means** | | | | | | | **Type III Tests of Fixed Effects** | |
| **FS measures** | **AD (n=26)** | **SCZ (n=89)** | **BD (n=116)** | **OCD (n=98)** | **SUD (n=120)** | **HC (n=145)** |  |  |
|  | **Mean (SE)** | **Mean (SE)** | **Mean (SE)** | **Mean (SE)** | **Mean (SE)** | **Mean (SE)** | **F** | **p_cor_** |
| Fixation frequency | 7.40(0.72) | 7.31(0.36) | 7.37(0.32) | 6.9(0.35) | 6.45(0.33) | 6.36(0.29) | 1.8 | 0.2 |
| Median fixation duration | 1028.41(219.74) | 919.95(110.30) | 899.51(97.85) | 905.30(107.20) | 1078.53(102.38) | 1186.62(87.48) | 1.49 | 0.2 |
| Saccade frequency | 5.04(0.7) | 5.17(0.38) | 4.96(0.3) | 4.56(0.32) | 3.97(0.31) | 4.07(0.27) | 2.44 | 0.2 |
| Mean saccade amplitude | 0.76(0.09) | 0.71(0.04) | 0.65(0.04) | 0.68(0.04) | 0.64(0.04) | 0.62(0.03) | 0.91 | 0.5 |
| Mean scanpath length | 4.61(0.69) | 4.08(0.34) | 3.72(0.3) | 3.64(0.33) | 3.22(0.32) | 3.09(0.27) | 1.76 | 0.2 |
| AD: Alzheimer’s dementia, BD: bipolar disorder, FS: fixation stability, HC: healthy control, p_cor_ : Adjusted p-value, OCD: obsessive-compulsive disorder, SCZ: schizophrenia, SE: standard error, SUD: substance (alcohol) use disorder | | | | | | | | |

| **Supplementary Table S8: Effects of age and sex on FS measures for trials without and with distractors when compared between AD, SCZ, BD, OCD, SUD and HC** | | | | | | | | | | |
| --- | --- | --- | --- | --- | --- | --- | --- | --- | --- | --- |
|  | **Fixation frequency** | | **Median duration of fixation** | | **Saccade frequency** | | **Mean saccade amplitude** | | **Mean scanpath length** | |
|  | **F** | **p_cor_** | **F** | **p_cor_** | **F** | **p_cor_** | **F** | **p_cor_** | **F** | **p_cor_** |
| **For trials without distractors** | | | | | | | | | | |
| **Intercept** | 116.335 | <0.001 | 122.973 | <0.001 | 32.879 | <0.001 | 110.547 | <0.001 | 24.306 | <0.001 |
| **Diagnosis** | 1.802 | .2 | 1.492 | .2 | 2.442 | .2 | .908 | .5 | 1.757 | .2 |
| **Sex** | 1.631 | .34 | .895 | .34 | 1.050 | .34 | 1.152 | .34 | 2.437 | .34 |
| **Age** | 19.414 | <0.001 | 24.921 | <0.001 | 27.978 | <0.001 | 2.194 | .14 | 14.119 | <0.001 |
| **For trials with distractors** | | | | | | | | | | |
| **Intercept** | 153.739 | <0.001 | 152.391 | <0.001 | 52.572 | <0.001 | 196.923 | <0.001 | 35.665 | <0.001 |
| **Diagnosis** | 7.331 | <0.001 | 4.526 | <0.001 | 7.342 | <0.001 | 4.100 | .001 | 7.792 | <0.001 |
| **Sex** | 1.096 | .7 | 2.995 | .4 | .586 | .7 | .084 | .8 | .059 | .8 |
| **Age** | 37.936 | <0.001 | 26.982 | <0.001 | 47.144 | <0.001 | 17.683 | <0.001 | 33.296 | <0.001 |
| FS: fixation stability, FDR: first-degree relatives, HC: healthy control, p_cor_ : Adjusted p-value | | | | | | | | | | |

| **Supplementary Table S9: Group comparisons between SCZ, BD, OCD, SUD and HC for trials with distractors** | | | | | | | | |
| --- | --- | --- | --- | --- | --- | --- | --- | --- |
| **Estimated marginal means** | | | | | | **Type III Tests of Fixed Effects** | | |
| **FS measures** | **SCZ (n=89)** | **BD (n=116)** | **OCD (n=98)** | **SUD (n=120)** | **HC (n=145)** |  |  |  |
|  | **Mean (SE)** | **Mean (SE)** | **Mean (SE)** | **Mean (SE)** | **Mean (SE)** | **F** | **p_cor_** | **Posthoc** |
| Fixation frequency | 8.12(0.32) | 8.36(0.29) | 7.25(0.32) | 6.39(0.3) | 6.68(0.25) | 8.69 | <0.001 | SCZ,BD>HC, SUD |
| Median fixation duration | 684.32(83.58) | 720.70(74.14) | 889.58(80.97) | 1048.3(78.04) | 1057.46(65.58) | 5.39 | <0.001 | SCZ, BD<HC, SUD |
| Saccade frequency | 5.88(0.31) | 6.12(0.27) | 4.91(0.3) | 4.33(0.29) | 4.47(0.24) | 8.72 | <0.001 | SCZ>HC,SUD;  BD>HC,SUD,OCD |
| Mean saccade amplitude | 0.96(0.04) | 0.97(0.04) | 0.83(0.04) | 0.81(0.04) | 0.82(0.03) | 4.94 | 0.001 | SCZ, BD>HC,SUD |
| Mean scanpath length | 6.23(0.39) | 6.62(0.35) | 4.7(0.38) | 4.37(0.36) | 4.51(0.3) | 8.89 | <0.001 | SCZ,BD>HC,SUD,OCD |
| AD: Alzheimer’s dementia, BD: bipolar disorder, FS: fixation stability, HC: healthy control, p_cor_ : Adjusted p-value, OCD: obsessive-compulsive disorder, SCZ: schizophrenia, SE: standard error, SUD: substance (alcohol) use disorder | | | | | | | | |

| **Supplementary Table S10: Effects of age and sex on FS measures for trials with distractors when compared between SCZ, BD, OCD, SUD and HC groups** | | | | | | | | | | |
| --- | --- | --- | --- | --- | --- | --- | --- | --- | --- | --- |
|  | **Fixation frequency** | | **Median duration of fixation** | | **Saccade frequency** | | **Mean saccade amplitude** | | **Mean scanpath length** | |
|  | **F** | **p_cor_** | **F** | **p_cor_** | **F** | **p_cor_** | **F** | **p_cor_** | **F** | **p_cor_** |
| **Intercept** | 177.009 | <0.001 | 185.810 | <0.001 | 56.541 | <0.001 | 232.588 | <0.001 | 34.794 | <0.001 |
| **Diagnosis** | 8.685 | <0.001 | 5.394 | <0.001 | 8.719 | <0.001 | 4.943 | .001 | 8.894 | <0.001 |
| **Sex** | 1.452 | .5 | 3.125 | .4 | .757 | .64 | .102 | .88 | .025 | .88 |
| **Age** | 41.371 | <0.001 | 26.938 | <0.001 | 50.977 | <0.001 | 18.012 | <0.001 | 37.124 | <0.001 |
| FS: fixation stability, FDR: first-degree relatives, HC: healthy control, p_cor_ : Adjusted p-value | | | | | | | | | | |

| **Supplementary Table S11: Model fit as per Bayesian information criteria scores for trials with and without distractors when compared between affected groups** | | | | | | | | | |
| --- | --- | --- | --- | --- | --- | --- | --- | --- | --- |
| **AD vs SCZ vs BD vs OCD vs SUD vs HC** | | | | | | | **SCZ vs BD vs OCD vs SUD vs HC** | | |
|  | **Trials without distractors** | | | **Trials with distractors** | | | **Trials with distractors** | | |
|  | **Age, sex & diagnosis** | **Age & sex** | **Diagnosis & sex** | **Age, sex & diagnosis** | **Age & sex** | **Diagnosis & sex** | **Age, sex & diagnosis** | **Age & sex** | **Diagnosis & sex** |
| **Fixation frequency** | 3135.34 | 3145.62 | 3147.23 | 3007.68 | 3043.45 | 3036.97 | 2867.20 | 2900.06 | 2899.62 |
| **Median fixation duration** | 9846.74 | 9912.73 | 9875.40 | 9501.01 | 9578.96 | 9531.07 | 9111.80 | 9176.74 | 9141.84 |
| **Saccade frequency** | 3069.04 | 3081.81 | 3089.07 | 2955.11 | 2990.46 | 2992.98 | 2813.25 | 2845.80 | 2854.56 |
| **Mean saccade amplitude** | 649.16 | 633.69 | 639.86 | 500.01 | 499.10 | 505.72 | 487.24 | 488.93 | 493.28 |
| **Mean scanpath length** | 3084.75 | 3094.31 | 3091.36 | 3227.14 | 3267.10 | 3252.50 | 3070.95 | 3106.08 | 3099.83 |
| AD: Alzheimer’s dementia, FS: fixation stability, FDR: first-degree relatives, HC: healthy control | | | | | | |  |  |  |

| **Supplementary Table S12: Partial correlation of FS performance measures and CGI severity scores in the affected group** | | | | | | |
| --- | --- | --- | --- | --- | --- | --- |
|  | **Trials without distractors** | | | **Trials with distractors** | | |
| **FS measures** | r | p | p_cor_ | r | p | p_cor_ |
| Fixation frequency | -0.05 | 0.27 | 0.45 | -0.15 | 0.004 | 0.006* |
| Median fixation duration | -0.03 | 0.57 | 0.71 | 0.1 | 0.05 | 0.06 |
| Saccade frequency | -0.09 | 0.06 | 0.3 | -0.16 | 0.002 | 0.006* |
| Mean saccade amplitude | 0.01 | 0.88 | 0.88 | -0.09 | 0.09 | 0.09 |
| Mean scanpath length | -0.06 | 0.24 | 0.45 | -0.15 | 0.003 | 0.006* |
| CGI: Clinical Global Impression, FS: fixation stability, p_cor_ : Adjusted p-value | | | | | | |

| **Supplementary Table S13: Group comparisons between SCZ, BD vs FDRs of SCZ/BD vs HC for trials with distractors** | | | | | | |
| --- | --- | --- | --- | --- | --- | --- |
|  | **Estimated marginal means** | | | **Type III Tests of Fixed Effects** | | |
| **FS measures** | **Affected (n=201)** | **FDR (n=237)** | **HC (n=145)** |  |  |  |
|  | **Mean (SE)** | **Mean (SE)** | **Mean (SE)** | **F** | **p_cor_** | **Posthoc** |
| Fixation frequency | 8.33 (0.22) | 6.79 (0.21) | 6.74(0.25) | 18.42 | <0.001 | Affected > FDR, HC |
| Median fixation duration | 693.66 (53.95) | 979.52 (50.43) | 1035.7 (63.06) | 11.43 | <0.001 | Affected < FDR, HC |
| Saccade frequency | 6.08 (0.21) | 4.59 (0.2) | 4.52 (0.24) | 19.24 | <0.001 | Affected > FDR, HC |
| Mean saccade amplitude | 0.97 (0.03) | 0.86 (0.02) | 0.82 (0.03) | 9.66 | <0.001 | Affected > FDR, HC |
| Mean scanpath length | 6.53 (0.27) | 4.86 (0.25) | 4.58 (0.31) | 16.49 | <0.001 | Affected > FDR, HC |
| FS: fixation stability, FDR: first-degree relatives, HC: healthy control, SCZ: schizophrenia, SE: standard error, p_cor_ : Adjusted p-value | | | | | | |
